# Experiences towards hormonal treatments: a qualitative study among endometriosis patients and healthcare professionals

**DOI:** 10.64898/2026.03.31.26349847

**Authors:** Domitille Le Quéré, Manon Verroul, Agathe Thierry, Mathilde Bouvard, Emma Brault-Galland, Gil Dubernard, Charles-André Philip, Julie Haesebaert, Axelle Brulport

## Abstract

**Objective:** To investigate, in the context of endometriosis management, the perceptions of patients and healthcare professionals regarding hormonal treatment options.

**Design:** Qualitative study using semi-structured focus group methodology.

**Setting:** University hospitals and academic research center.

**Subject(s):** Patients with endometriosis (n=20) and healthcare professionals (n=13) involved in their care.

**Intervention(s):** Not applicable

**Main Outcome Measure(s):** Focus group topics investigated representations on the concept of treatment effectiveness, emotion associated to this medical management and the perceived impact of these therapies on patient-professional and patient-environment relationship.

**Result(s):** We highlighted a discrepancy between patients and doctors regarding the concept of efficacy of hormonal therapies. Long-term amenorrhea is the main priority for healthcare professionals, whereas pain reduction remains the immediate wait for patients. Interviewed patients reported a lack of listening and empathy, a shared-information deficit as regards treatment options and side-effects and a need to involved partner and family in care. These factors contribute to communication issues between patients and doctors and appear to contribute to significant mental burden on both sides. Among healthcare professionals, mental burden appears to arise primarily from the resource-intensive demands of endometriosis management, whereas among patients it is driven more by the need to take an active role in their own care to compensate for insufficient information provided by physicians.

**Conclusion:** In this study, we highlighted the ambiguities surrounding the concept of therapeutic efficacy of hormonal therapies and collected several factors to try to improve shared-decision-making process in the management of endometriosis. This is designed to help us create a shared decision-making tool in the near future.

## INTRODUCTION

Endometriosis is a chronic gynecological disease characterized by endometrial-like tissue outside the uterus. Usually, the development of this ectopic tissue is associated with oestrogen-dependent, proinflammatory, proangiogenic, immunologic and neurogenesis processes that appear to be interdependent and leading to a variety of symptoms. Their heterogeneity includes infertility, fatigue, depression, heavy menstrual bleeding and a wide range of painful symptoms (pelvic pain, dyspareunia, dysmenorrhea, painful ovulation, dysuria and functional bowel disorders) (1,2). Due to these clinical manifestations, endometriosis is a debilitating disease associated with a significant impairment of the overall women’s quality of life (QoL), mental health and work productivity (3,4). The annual economic burden of endometriosis, including health care and productivity loss, was estimated to be €10,000 per patient in Europe and $27,855 in the US (5,6).

Currently, there is no cure for endometriosis. The palliative treatment options for endometriosis-associated painful symptoms include medical treatments, non-pharmacological treatments and surgery (1,2,7). Among medical therapies, hormonal treatments are strongly recommended to treat pain symptoms and are the first-line-therapeutic option for patients (7,8). Many negative side effects are related to the hormonal therapies and these treatments are not an option for patients who wish to become pregnant (2). Probably related to central sensitization or a physiopathological mechanism of progesterone resistance, between 10% and 30% of women with endometriosis have no pain reduction with hormonal medications (1,2,9). To date, there is no way of predicting patient response to available hormone therapies in order to propose a suitable treatment and limit therapeutic failures and treatment non-compliance.

These concerns help explain why endocrine therapies are increasingly unpopular among women with endometriosis. In their survey of 1,420 patients from Germany, Austria, and Switzerland, Burla et al. highlighted negative experiences with hormonal pills, particularly regarding side-effects, changes in libido, disruption of hormonal and menstrual cycles, insufficient efficacy, and the burden of daily intake (10). Thrombosis, osteoporosis, psychological effects, weight gain, bloating, fear of cancer and skin blemishes complete this list of reported negative experiences (11). These experiences contribute to negative mental representations (side effects, pain, inneffective, depression, anxiety, uncertainty) associated with hormonal endometriosis therapies (11). The use of hormonal treatments is often linked to a lack of alternatives, and non-endocrine drugs would be preferred by women with endometriosis (10,11). Several drug innovation and repositioning are under clinical investigation for endometriosis (2,12). None of them has yet identified a non-endocrine compounds really effective in endometriosis (12). Given our knowledge of the pathophysiology of endometriosis, it is still difficult to say whether a non-endocrine drug or surgery alone could allow patients to avoid hormonal therapies in the management of endometriosis.

Endometriosis requires thus complex and repeated treatment decisions, yet patient engagement and patient-professional relationship is often undermined by limited evidence about hormonal treatments choices and inconsistent communication (13,14). Despite international guidelines and patients representatives calling for patient engagement in decisions notably through shared decision-making, little research or decision aids focused on hormonal therapies—the cornerstone of long-term management (7,13,15,16). The aim of this cross-sectional qualitative study is to clarify how patients and healthcare professionals perceive the efficacy of these treatments given their own priority in order to identify ways to strengthen therapeutic alliances and improve quality and experience of care.

## MATERIALS AND METHODS

### Study design

We conducted a qualitative study using focus groups with women living with endometriosis and healthcare professionals involved in their care. Focus groups were selected to explore social representations of hormone treatments and the perception of endometriosis impact on women’s lives. Often used for health research, this methodology allows for exploration of collective representations and experiences of healthcare system users (17). Focus groups have proven to be a particularly relevant method for conducting studies with several populations in parallel (e.g., caregivers, patients), enabling researchers to “grasp the full psychosocial complexity of health issues”. Sensitive topics can be encouraged by the dynamics of a group approach, but also by the presence of similarities between members who share a common experience and identity (17,18). Results are reported according to the Standards for Reporting Qualitative Research guidelines (19). The study received ethics approval from the Hospices Civils de Lyon ethics committee, and personal data were handled in conformity with the general data protection regulation.

### Patient and Public Involvement (PPI)

In line with the recommendations emphasizing the central role of PPI in producing relevant and acceptable health research, the study was co-developed with a patient partner. We reported her implication in the study based on GRIPP2 short form reporting checklist (20). A woman with lived experience of endometriosis, trained in patient–professional partnership acted as co-investigator. She was part of the steering committee and contributed to study design, data collection, analysis, interpretation, and dissemination. She was involved in interview guides development and co-facilitated patient focus groups. Patient participants were also invited to contribute beyond data collection through the organization of a focus groups dedicated to validation and interpretation of findings and prioritization of future research. Financial compensation was provided to acknowledge patient partner and participants’ contribution based on the time devoted to the study.

### Participants and recruitment

For patients, the inclusion criteria were being between 18–50 years; confirmed diagnosis of endometriosis (imaging or surgery); experience of at least three lines of hormonal therapy (oral contraceptives, hormonal IUD, dienogest, GnRH analogues) and available for two focus groups. Women in physiological menopause were not included. Women were recruited between June and December 2024 via two main French patient associations approved by the French Ministry of Health and through 2 gynecology departments from the second largest French teaching hospital. The two referral centers for endometriosis also participated to patents’ recruitment. The call for participation was also shared on social media.

The healthcare professionals were gynecologists or midwives who devoted at least 30% of their practice to treating endometriosis. Professionals were recruited between August 2024 and January 2025 in two regionals networks involving professionals specializing in care of endometriosis and among trainees from a national university diploma on endometriosis management.

All participants, patients and professionals, gave their consent to participation and audiorecording of the interviews.

### Interview guide and data collection

Each group of patient consisted of between 6 and 10 participants attending in person. Although they took place at the Hospices Civils de Lyon, patients were invited to a neutral location outside gynecological services. To structure the focus groups, an interview guide was developed based on existing literature and experiential knowledge of steering committee members. It was organized in four topics, each introduced with a visual aid: (1) perceived treatment effectiveness with a scale of effectiveness; (2) the therapeutic journey addressed using an emotion wheel to describe their experiences; (3) the patient–professional relationship introduced with a metaplan; (4) the patient–environment relationship, evoked through photolanguage. Where necessary, follow-up questions were provided to explore the themes in greater depth. The interview guide is presented in Supplementary Table S1. After the focus groups, a second meeting was offered to all study participants and the research team. It consisted of a presentation of the results and an open discussion on the participants’ perceptions of the relevance and completeness of the results, their understanding, and their implications for future research. This exchange took place online using Microsoft Teams to allow as many participants as possible to attend.

For professionals, each group included 6–10 participants. Given the limited availability of professionals, the groups met exclusively online using Microsoft Teams. They were co-facilitated by the psychologist and a pharmacy resident. Interview guide was also developed based on existing literature and experiential knowledge of steering committee members. It addressed the same topics as from the patient but with different materials: (1) the caregiver-patient relationship introduced with a picture; (2) the effectiveness of treatments with a gradient; (3) the therapeutic pathway, with a timeline; (4) the patient-environment relationship, addressed with Wooclap. The interview guide is presented in Supplementary Table S2.

### Data analysis

All discussions were audio-recorded, transcribed verbatim, and supplemented by field notes. Thematic analysis followed Braun and Clarke’s inductive framework (21). The first stage, coding, transforms raw data into meaningful units that can be used to identify characteristics that meet the objectives of the qualitative survey. Two to three researchers double-blind coded all transcripts to limit bias and ensure methodological rigor. Coding was performed on Taguette (22). The codes obtained were then sorted and organized into themes and sub-themes with the psychologist and discussed with the patient partner and other researchers. Data saturation was assessed iteratively (23). The content of the interviews with patients and professionals was analyzed independently. The results were then compared to identify differences and similarities between the two populations. The results were shared with the participants in a dedicated online discussion group, to which all participants from the first phase were invited. The final analysis incorporated the views and critical review of all members of the steering committee and participating women.

## RESULTS

### Patient’s perception of hormonal treatments in the management of endometriosis

We conducted three focus group including twenty patients (mean age 34.9 ± 7 years) (Table 1). All of them have been diagnosed with endometriosis through imaging or surgery, and 90% have or have had clearly identified lesions. The majority of patients experienced chronic pelvic pain (75%), digestive symptoms (70%), and dysmenorrhea (55%). Most participants showed signs of pain sensitivity (55%). The average of the tested hormonal treatment lines was 5.1 ± 1.3. The main reasons for discontinuation were multiple intolerance, persistent pain and bleeding. Four major themes emerged from the focus group analyses (Table 3).

**Table 1.** Baseline characteristics of patient cohort.

|  | Total cohort<br>(n=20) | Focus<br>group 1<br>(n=5) | Focus group<br>2 (n=9) | Focus<br>group 3<br>(n=6) |
| --- | --- | --- | --- | --- |
| Age - Mean (SD) | 34.9 ± 7.0 | 36.2 ± 7.4 | 35.8 ± 6.8 | 32.3 ± 7.4 |
| Marital status - n(%) |  |  |  |  |
| Married, committed, relationship | 12 (60) | 5 | 5 | 2 |
| Single | 8 (40) | 0 | 4 | 4 |
| Parity - n(%) |  |  |  |  |
| Yes | 4 (20) | 1 | 1 | 2 |
| No | 16 (80) | 4 | 8 | 4 |
| Fertility problems - n(%) |  |  |  |  |
| Yes | 9 (45) | 2 | 4 | 3 |
| No | 6 (30) | 1 | 2 | 3 |
| Unknown | 5 (25) | 2 | 3 | 0 |
| Diagnosis delay (years) - Mean (SD) | 7.4 ± 8.4 | 4.8 ± 6.8 | 11.0 ± 10.1 | 3.5 ± 3.1 |
| Diagnostic methods - n(%) |  |  |  |  |
| Ultrasound and/or MRI | 17 (85) | 4 | 8 | 5 |
| Surgery | 3 (15) | 1 | 1 | 1 |
| Macroscopic lesions (US, MRI and/or surgery) - n(%) |  |  |  |  |
| Yes | 18 (90) | 4 | 8 | 6 |
| No | 1 (5) | 0 | 1 | 0 |
| Unknown | 1 (5) | 1 | 0 | 0 |
| Symptoms - n(%) |  |  |  |  |
| Chronic pelvic pain | 15 (75) | 4 | 7 | 4 |
| Digestive symptoms | 14 (70) | 4 | 9 | 1 |
| Dysmenorrhea | 11 (55) | 3 | 5 | 3 |
| Dyspareunia | 8 (40) | 1 | 4 | 3 |
| Urinary symptoms | 7 (35) | 2 | 3 | 2 |
| Asthenia | 7 (35) | 1 | 3 | 3 |
| Mood disorders | 3 (15) | 0 | 3 | 0 |
| Others: low back, scapula and chest pain, neuropathic pain and dyspnea | 4 (20) | 0 | 2 | 2 |
| Pelvic pain sensitization - Convergences PP score () - n(%) |  |  |  |  |
| Positive (≥ 5) | 11 (55) | 2 | 6 | 3 |
| Negative (< 5) | 9 (45) | 3 | 3 | 3 |
| Surgery - n(%) |  |  |  |  |
| Yes | 11 (55) | 3 | 5 | 3 |
| No | 9 (45) | 2 | 4 | 3 |
| Mean number of hormonal treatment lines per patient - Mean (SD) | 5.1 ± 1.3 | 5.4 ± 1.1 | 5.2 ± 1.6 | 4.7 ± 1.0 |
| Treatment received at least once - n(%) |  |  |  |  |
| Dienogest | 15 (75) | 5 | 6 | 4 |
| Ethinylestradiol (EE) + Levonorgestrel | 14 (70) | 3 | 6 | 5 |
| Low-dose progestin therapy | 14 (70) | 4 | 6 | 4 |
| GnRH agonists | 8 (40) | 1 | 5 | 2 |
| High-dose progestin therapy | 7 (35) | 2 | 4 | 1 |
| EE + Cyproterone acetate | 4 (20) | 1 | 2 | 1 |
| Other combined estrogen-progestin therapy<sup>1</sup> | 5 (25) | 2 | 1 | 2 |
| Other hormonal treatments | 2 (10) | 0 | 1 | 1 |
| Cause of treatment discontinuation - n |  |  |  |  |
| Treatment intolerance<sup>2</sup> | 40 | 12 | 15 | 13 |
| Persistent pain | 29 | 1 | 14 | 14 |
| Bleeding | 20 | 4 | 9 | 7 |
| Change of gynecologist/midwife | 12 | 2 | 4 | 6 |
| Desire for pregnancy | 8 | 3 | 3 | 2 |
| Others<sup>3</sup> | 7 | 1 | 4 | 2 |

<sup>1</sup> EE + Dienogest, Drospirenone, Nomegestrol, Norgestimate or Gestodene
<sup>2</sup> headaches (experienced 10 times), weight gain (10), acne (8), depression (7), digestive symptoms (5), libido disorders (5), limb oedema (4), insomnia (2), thoracic symptoms, arthralgias, asthenia, recurrent mycosis and ovarian cyst (experienced one time for each)
<sup>3</sup> meningioma (experienced 4 times), history of deep vein thrombosis (2), the will of changing the contraceptive method (4), temporary post-operative treatment (1) and end of marketing (1)

**Table 2.**
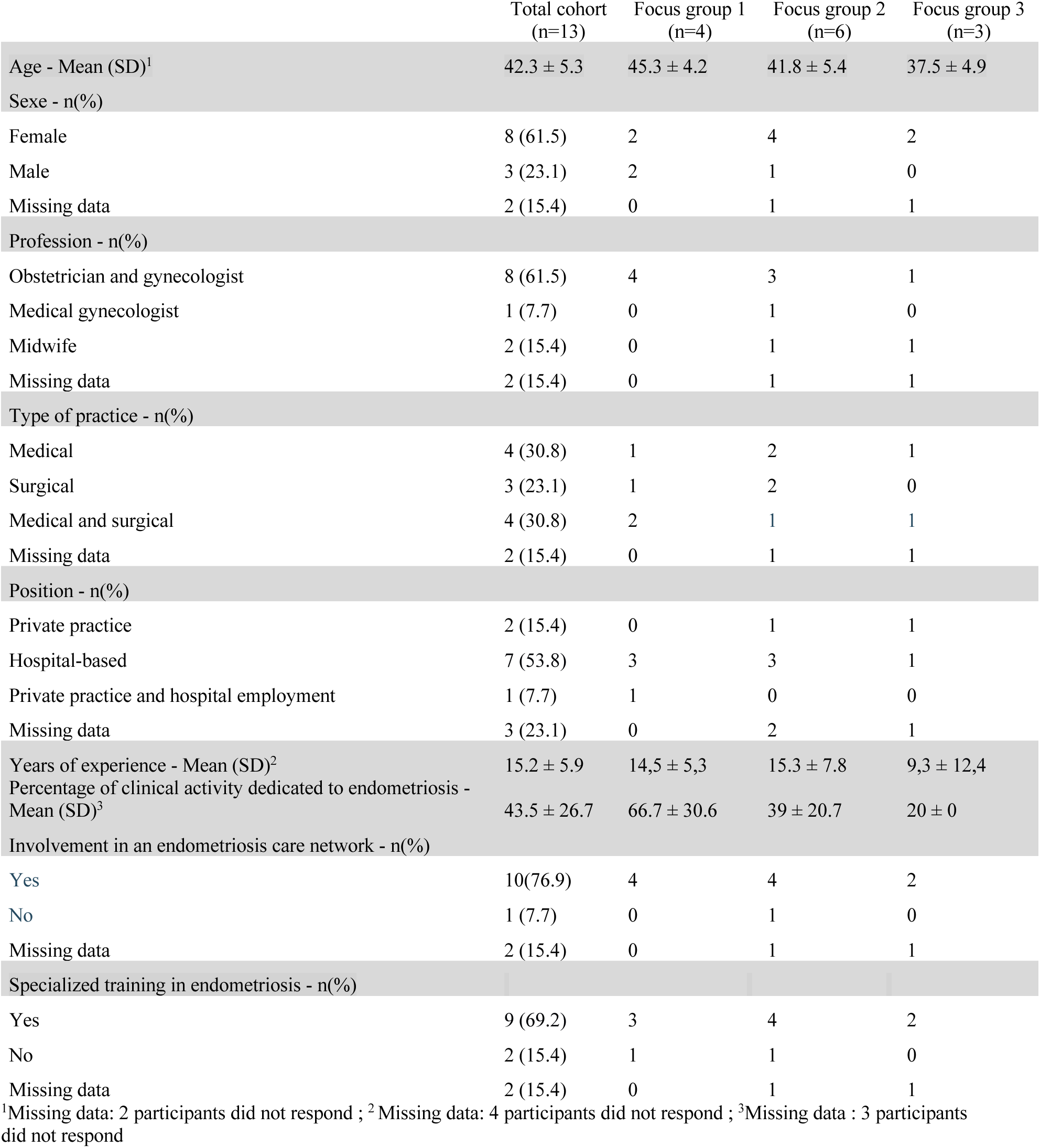
Baseline characteristics of the healthcare professional cohort.

**Table 3:**
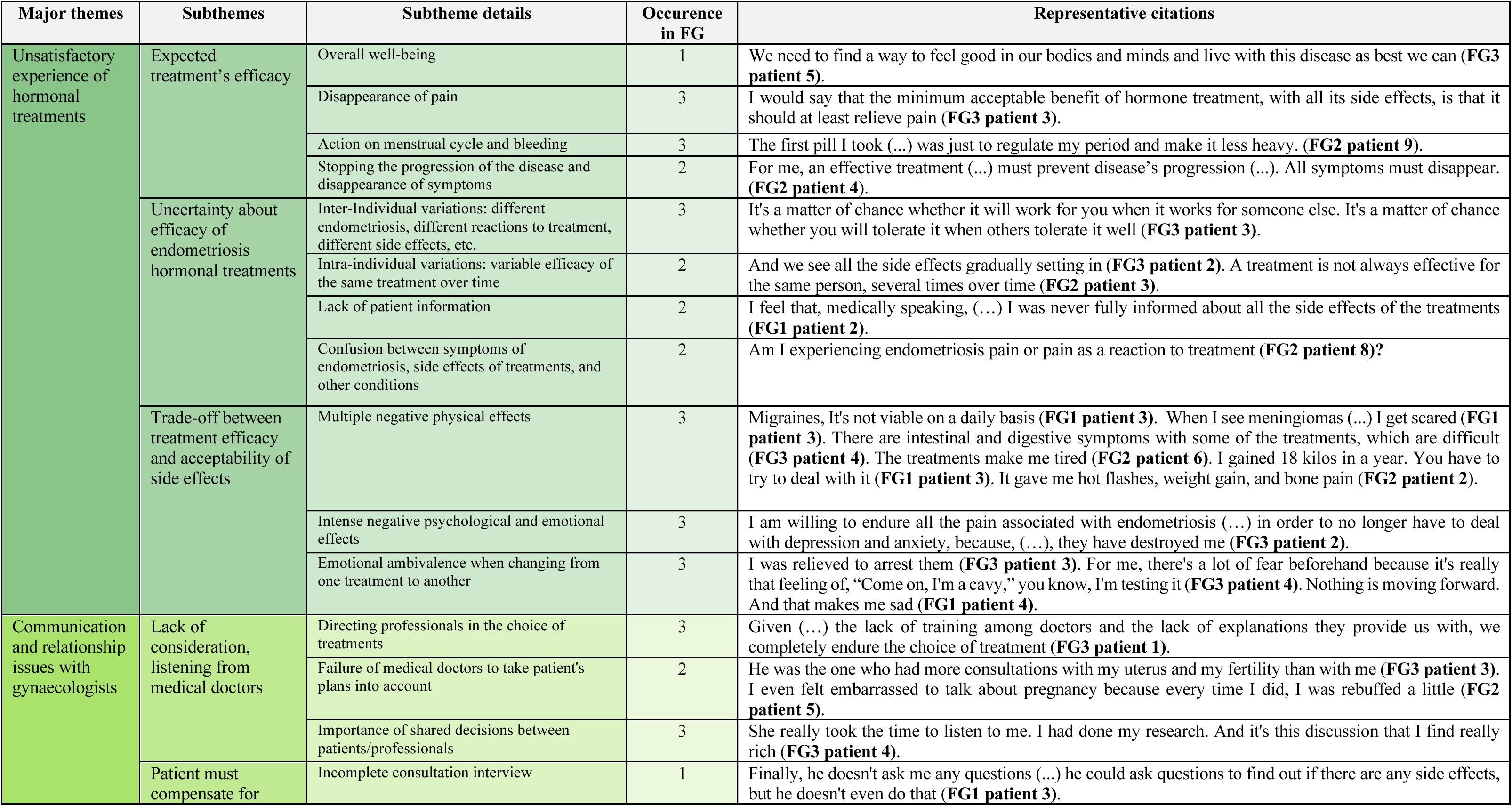
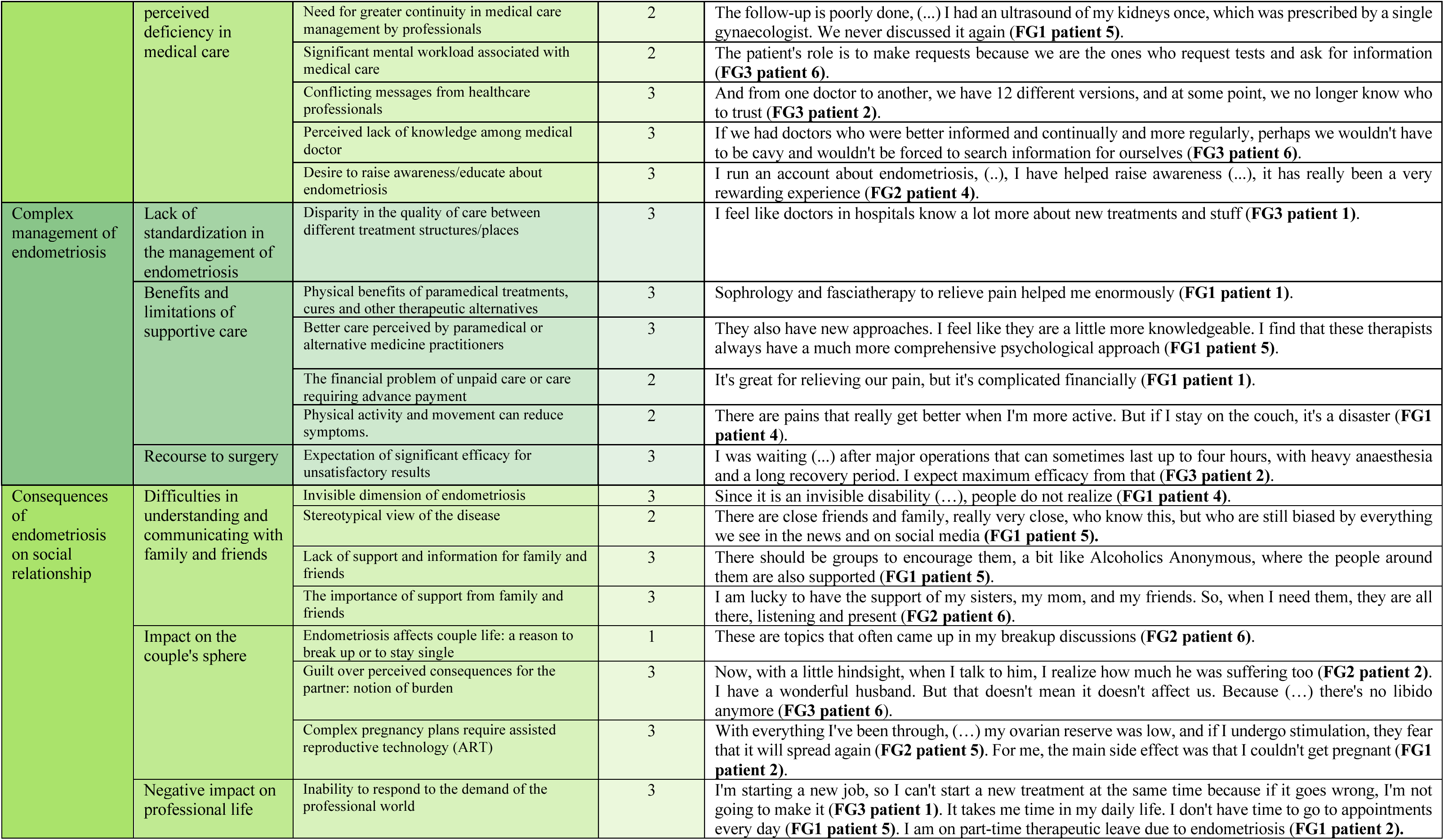
Patient’s perception of hormonal treatments in the management of endometriosis.

#### Major theme 1: Perceptions and Limitations of Hormonal Treatment Efficacy (Table 3)

Expected efficacy from hormonal treatment for our patients was first to limit pain, at a lesser extent to regulate menstrual flow, and for some, ideally, halt the disease’s progress. The many sides effect physical, psychological and emotional make essential to find a tradeoff between the expected benefits of treatment and its side effects. The decision between several treatment options is made difficult due to the lack of information by medical professional and the uncertainty about efficacy and side effect. Patients describe significant interindividual and individual variation of treatment efficacy over time but also confusion between the symptoms associated with endometriosis and the side effects of treatment. Therefore, patients often perceive themselves as being experimented on during treatment change, leading to anxiety and fear. Treatment changes seem to generate emotional ambivalence between the fear of the comeback of the pain and the relief to halt the last treatment. The hope of symptom improvement with treatment is often overshadowed by disappointment at the lack of real progress.

#### Major theme 2: Communication and relationship issues with gynaecologists (Table 3)

Patients emphasize importance of endometriosis care based on shared decisions with healthcare professionals. However, they report a lack of listening and consideration from physician, who often adopt a directive approach in treatment’s choices without taking sufficient account of their expectations and projects. They report feeling subjected to treatment decisions without having received the necessary information about side effects and long-term consequences. It generates a feeling of anger towards the medical community. Perceived lack of knowledge, inconsistent messages and incomplete consultation interview from healthcare professionals, lead to a feeling of failure of the medical community. Patient believe that they are expected to compensate these failures by keeping themselves informed and assuring the continuity of care, which is experienced as significant mental burden.

#### Major theme 3: Complex management of endometriosis (Table 3)

Patients described endometriosis management as involving many different actors. They perceived that differences in healthcare structures and regional disparities resulted in unequal quality of care, depending on the facilities available to them. Many reported that support care, alternative medicine, and physical activity contributed substantially to symptom relief. In addition, patients report a better experience with paramedical and alternative medicine, where professionals are more attentive and better informed. However, the lack of reimbursement for this care in France seems to be a factor that limits access for all patients. Patients explained that when surgery was proposed after insufficient response to medical therapy, they placed great expectations on its ability to relieve symptoms. Yet many expressed disappointments, pointing to side effects, the continuation of hormonal treatment, and persistent pain.

#### Major theme 4: Consequences of endometriosis on social relationships (Table 3)

Relationship issues with family and friends were reported to be related to the invisible nature of the disease, the stereotypical view of endometriosis conveyed by social media, and the lack of support from relatives in their role as caregivers. These realities led patient to feel lonely. Therefore, patients give great importance on raising awareness and educating people about endometriosis, particularly among young people. In addition, many described how their couple’s life was affected by disrupted sexual life, infertility, and the perception of being a burden to their partner, which sometimes resulted in breakups or rejection of a shared life. These life situations were reported to be associated with feelings of loneliness and sadness. Endometriosis also affected negatively their professional life preventing them from meeting the demands of the workplace. Endometriosis may therefore require adjustments to be made in the workplace.

### Healthcare professional’s perceptions of patient’s experience of hormonal treatments in the management of endometriosis

Thirteen healthcare professionals participated in three dedicated focus group (Table 2). Baseline characteristics were collected with a self-reported questionnaire. The average age of the participants was 42.3 ± 5.3 years with an average experience of 15.2 ± 5.9 years. More than a half of participants were obstetricians and gynecologist (61.5%), worked at hospital (53.8%) and have a surgical practice. More than 76% were involved in endometriosis care network, and 69.2% received a specialized training in endometriosis.

#### Major theme 1: Relational issues and communication modality (table 4)

Professionals highlight the pivotal role of the first consultation. Each patient arrives with their own history, expectations and emotions, shaping the patient–provider relationship and care pathway. Professionals consider it essential to fully understand the complexity of each patient’s journey and to identify their expectations in order to provide appropriately tailored care. Maintaining a professional posture and building a relationship of trust are key. Professionals favor a collaborative over a hierarchical model, based on listening and sharing information, which supports treatment adherence and addresses patients’ negative emotions. Given their complexity and duration, consultations are described as long and energy-intensive.

**Table 4:**
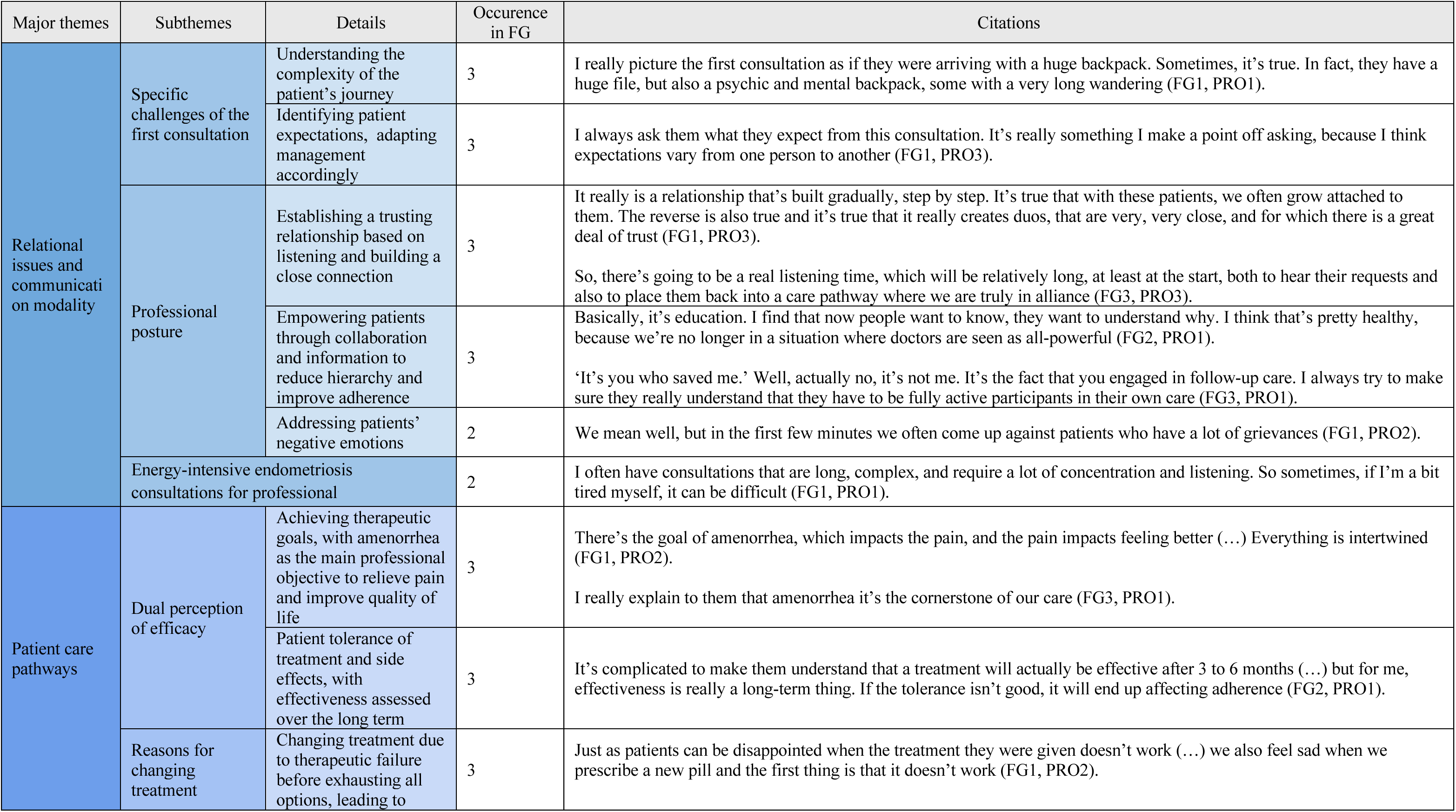
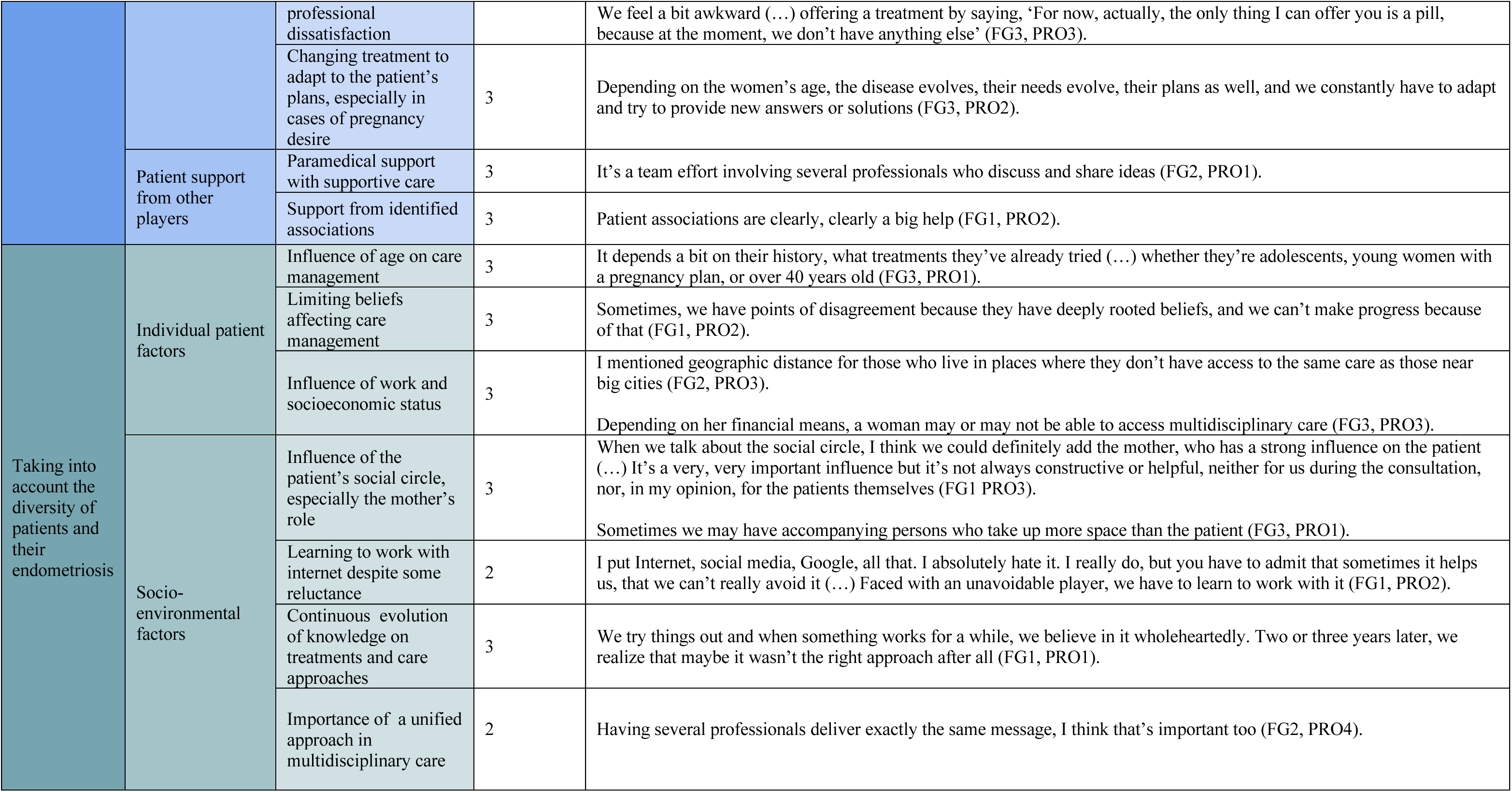
Healthcare professional’s perceptions of patient’s experience of hormonal treatments in the management of endometriosis.

#### Major theme 2: Patient care pathways (table 4)

For professionals, treatment effectiveness is defined by the achievement of amenorrhea over the long term, a mandatory precondition for improving pain symptoms and quality of life. When amenorrhea is achieved, treatment tolerance must be considered, since acceptable tolerability is a prerequisite for patient compliance. Except when predetermined objectives are not reached, the two main reasons prompt treatment changes for professionals are adapting to the patient’s plans, such as pregnancy, and therapeutic failure. Furthermore, despite multiple options offered by hormonal treatments, surgery, supportive and multidisciplinary care, a therapeutic impasse can occur leading to dissatisfaction among professionals. The support offered to patients through other professionals including physical therapist, pain specialist, psychologist and dieticians, or patient associations complements the ongoing care provided.

#### Major theme 3: Taking into account the diversity of patients and of their endometriosis (table 4)

According to the gynecologists interviewed, both individual and socio-environmental factors affect therapeutic pathways, communication and disease management. They reported that while age mainly influences communication and care choices, patient’s beliefs, socioeconomic status and social support network -for instance their mother- can influence negatively the management provided. Internet also plays a growing role as a source of information and influences patients, requiring professionals to guide patients toward reliable resources. Professionals reported that clinical practices and treatment knowledge are continuously evolving, requiring them to make ongoing adjustments. Finally, to their point of view, consistency and coherence within a multidisciplinary framework are essential to strengthen patient understanding, therapeutic adherence and counter limiting beliefs.

## DISCUSSION

In this qualitative study, based on focus group methods, we explored various aspects inherent to the use of hormonal therapies in the management of endometriosis. We reported here the central role of communication and analysed, at different levels, the factors underlying relational issues that affect patient care and quality of life. In this study, the involvement of a patient partner contributed to make sure that the research question and each stage of the study were approached with a view that was closely aligned with the lived experiences of patients. Her co-facilitation of the focus groups promoted the open expression of each participant. Her contribution to the analysis of the results ensured that patient testimonies were consistently interpreted within the broader context of the discussions, reducing the risk of misinterpretations.

### Doctor-patient communication issues concerning treatment strategies

We identified two major causes of communication issues between patients and healthcare professionals regarding the endometriosis management strategy with hormonal therapies.

The first concerns the definition of therapeutic efficacy. For healthcare professionals, the evaluation of treatment effectiveness is primarily based on amenorrhea, whereas for patients, pain reduction remains the main priority. It is interesting to note that hormonal treatment’s efficacy is perceived by both parties as uncertain and variable depending on the individual but that the explanations for these inter-individual variations differ depending on the viewpoints. For patient, this variability in treatment response is mainly attributed to personal factors. In contrast, healthcare professionals also highlighted the major influence of social environment, including family, social media and socio-economic conditions, on patient’s experience of illness and care.

The second relates to the time scale. For doctors, achieving long-term amenorrhea is a prerequisite for improving other symptoms. For their part, patients adopted an immediate perspective, viewing pain reduction as the essential condition for improving their quality of life. Patients place particular emphasis on the fact that these ambitious goals must not be achieved at the expense of their psychological well-being. Healthcare professionals are well aware that, with current treatment options, it is often difficult to achieve long-term amenorrhea associated with good tolerance of hormonal treatments. Even with the development of multidisciplinary care, it is not uncommon for professionals to find themselves at a therapeutic impasse with their patients with endometriosis.

According to the ESHRE guidelines, hormone therapies are strongly recommended to reduce endometriosis-associated pain and related symptoms, but the concept of long-term treatment and amenorrhea is not clearly addressed (7). This study, conducted among patients with long-term experience of hormonal management for endometriosis and clinicians with recognised expertise in the condition, reveals the necessity to revise the expected therapeutic aims of hormonal treatments using a timeline that reflects the realities of chronic disease care. Setting common objectives is key to improving the patient-professional relationship, especially since negative impression of the medical profession among women with endometriosis has already been associated with treatment failure (24).

### Doctor-patient relationship issues

There is a discrepancy between what doctors describe implementing in their practice and patient’s experiences. In our study, healthcare professionals declared themselves in favour of shared decision-making in the management of endometriosis, while patients found the patient-doctor relationship to be very directive and hierarchical. Patients reported a lack of attentive listening and consideration for their personal goals by gynecologists. They reported receiving more attentive listening from paramedical staff and expressed strong support for supportive care services. Patients with endometriosis would also like to be better informed about the different treatment options, particularly about the side effects and their short-, medium- and long-term consequences. It has already been reported that a shared-information deficit on the part of physicians, about treatment options and side effects in the context of endometriosis management, is a reality and a major obstacle in shared-decision-making process (11,13). In this study, from the patients’ perspective, this lack of listening and information reflects insufficient physician training and drives women to take an active role in managing their care by independently seeking information about the disease (through social media and patient associations). This process imposes a considerable mental burden and is not experienced as a form of empowerment, but rather as a necessity to achieve better health outcomes. For healthcare professionals, this directive approach may reflect insufficient training, difficulties in managing therapeutic dead ends, and/or an attempt to protect themselves from the emotionally and cognitively draining nature of consultations related to the complex management of endometriosis. Better clinician listening and training, greater empathy toward patients, giving full consideration to their reported symptoms, and involving them in decision-making have already been reported as important factors for improving the management of endometriosis (16,25).

### Patient-social relationship issues

The invisible nature of the disease, the stereotypes conveyed by social media, and the chronic and unpredictable aspects of the disease complicate relationships with those around them. Patients report that they sometimes end their social or professional relationships so that they do not have to justify themselves in order to avoid increasing the mental burden associated with the disease. They have expressed the need for support for those around them, because, as with all chronic diseases, the role of carers is essential. In ″ComPaRe-Endometriosis″ cohort, patients already reported the need of involving their partner or family in care (16).

### Study limitations

Several limitations merit consideration. First, as with any qualitative study utilizing purposive sampling, participants were volunteers who may have been more sensitized to endometriosis care issues than the broader target population. We sought to mitigate this selection bias by recruiting participants through diverse channels, thereby capturing a wider range of perspectives. Second, the healthcare professionals in our sample were practicing in referral centers and had received specialized training in endometriosis management, which may limit the generalizability of their perspectives to non-specialized providers. However, this potential bias likely skews toward more informed and patient-centered practices; consequently, we might expect the identified challenges and tensions between patients and professionals to be even more pronounced among non-specialized healthcare providers who lack dedicated endometriosis training. Third, our study included three focus groups each of patients and healthcare professionals. While this sample size is consistent with established qualitative methodologies for achieving data saturation, we acknowledge that additional groups might have revealed further nuances. Importantly, this represents the first study to directly compare and contrast the perspectives of patients and healthcare professionals regarding endometriosis care through parallel focus group methodology, providing novel insights into the relational dynamics within the therapeutic encounter.

## CONCLUSION

In this study, we highlighted i) the ambiguities surrounding the concept of therapeutic efficacy of hormonal therapies in the management of endometriosis and ii) some major obstacles in the implementation of shared-decision-making process. To improve endometriosis care, there is an urgent need to enhance communication quality between health care professionals and patients to reduce the mental burden of endometriosis and the negative emotions associated to the disease management. Our results advocates toward the development of a shared decision-support tool comprising dedicated components, i) for the various stakeholders involved in patient care (patients, clinicians, and relatives) including both scientific and educational content, ii) for the definition of a common therapeutic strategy with short-, medium-, and long-term objectives, and iii) for the formalization of red flags that would prompt a reassessment of the care plan.

## Data Availability

All data produced in the present study are available upon reasonable request to the authors

## CRediT Authorship Contribution Statement: Domitille Le Quéré

Formal analysis, Writing – original draft; **Manon Verroul**: Formal analysis, Methodology, Investigation, Writing – original draft; **Agathe Thierry**: Formal analysis, Investigation, Writing – original draft; **Mathilde Bouvard**: Formal analysis, Writing – original draft; **Emma Brault-Galland**: Investigation; **Gil Dubernard**: Conceptualization, Project administration; **Charles-André Philip**: Conceptualization, Project administration, Supervision, Validation, Visualization, Writing – original draft, Writing – review & editing; **Julie Haesebaert**: Conceptualization, Methodology, Project administration, Supervision, Validation, Visualization, Writing – original draft, Writing – review & editing; **Axelle Brulport**: Conceptualization, Methodology, Funding acquisition, Project administration, Supervision, Validation, Visualization, Writing – original draft, Writing – review & editing.

## Attestation statements

Data regarding any of the subjects in the study has not been previously published unless specified. Data will be made available to the editors of the journal pre and/or post publication for review or query upon request.

## Acknowledgments

We thank the patients for their time and effort in participating in this research. We are also very grateful to the two regional networks coordinating endometriosis care EndoAURA et EndoBFC and to Dr Giulia Gouy for helping us recruit the healthcare professionals in this study.

