## Supplementary Table S1 and S2 for "Experiences towards hormonal treatments: a qualitative study among endometriosis patients and healthcare professionals"

---

**<u>Table of content:</u>**

- Supplementary Table S1

- Supplementary Table S2

**Supplementary Table S1:** Patient focus group interview guide

| TOPICS | SUPPORT MATERIALS | QUESTIONS | FOLLOW-UPS |
|---|---|---|---|
| **Topic n°1 :**<br>**Perceived treatment effectiveness** | Scale of effectiveness<br>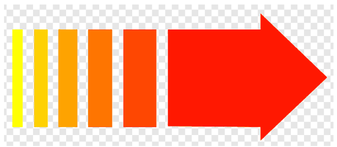 | *To understand your perception of the effectiveness of your treatment, we invite you to use the scale in front of you. It ranges from "minimal effectiveness" to "maximum effectiveness," and you can use it to answer the following question: How would you define the effectiveness of a treatment?* | - Concept of the temporality of effects (long term VS short term)<br>- Impact of treatment on the body and morale<br>- Possible negative effects: stress, anxiety, constraints |
| **Topic n°2 :**<br>**Therapeutic journey** | Emotion wheel<br>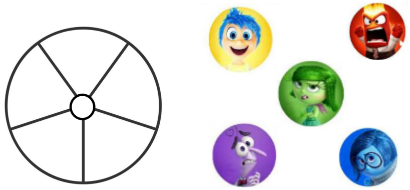 | *You have a circle at your disposal that we ask you to complete in order to best describe your experience during the changes in your therapeutic journey. You can use the emotions listed on the table (joy, fear, anger, sadness, disgust) or express yourself in your own words to explain how you experienced the different stages of the therapeutic journey ?* | - Choice of treatments<br>- Experience of changes<br>- Compromise between certain effects of one treatment and no other (constraints)<br>- Alternative/complementary medicine (in the experience of treatments) |
| **Topic n°3 :**<br>**The patient-professionnal relationship** | Métaplan<br>Professionnals \| Patients | *We now invite you to address the patient-healthcare professional relationship by completing the following table to describe, in your opinion, your role and that of your healthcare professionals in choosing your therapeutic treatments ?* | - Communication: explanations provided, understanding of changes (literacy)<br>- Shared decision-making: treatment chosen, overall vision, short- and long-term treatment management, life plan<br>- Various healthcare professionals consulted: their expectations, positioning, change of professional |
| **Topic n°4 :**<br>**The patiente-environnement relationship** | Photolanguage on the theme of social life | *We suggest we finish with your relationship with your social environment in the broadest sense. In front of you are some photos. We suggest you choose one or two images that represent the place your treatments occupy in managing your relationships with your environment. Afterwards, we will ask you to explain your choice, one at a time.* | - Communication<br>- Sources of information requested<br>- Social environment: relationship with partner, family life, caregiver/support person?<br>- Opinion of family and friends regarding treatment: importance given?<br>- Professional environment?<br>- Role of peers |

**<u>Supplementary Table S2:</u>** Healthcare professional focus group interview guide

| TOPICS | SUPPORT MATERIALS | QUESTIONS | FOLLOW-UPS |
|---|---|---|---|
| **<u>Topic n°1 :</u>** **The patient-professionnal relationship** | 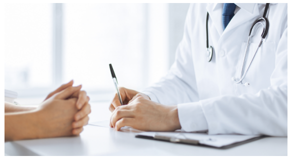 | *How would you describe your relationship with your patients throughout their treatment?* | - Concept of the temporality of effects (long term VS short term)<br>- Impact of treatment on the body and morale<br>- Possible negative effects: stress, anxiety, constraints |
| **<u>Topic n°2 :</u>** **Perceived treatment effectiveness** | 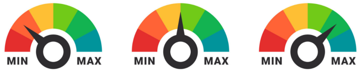 | *How would you define the effectiveness of a treatment?* | - Choice of treatments<br>- Experience of changes<br>- Compromise between certain effects of one treatment and no other (constraints)<br>- Alternative/complementary medicine (in the experience of treatments) |
| **<u>Topic n°3 :</u>** **Therapeutic journey** | 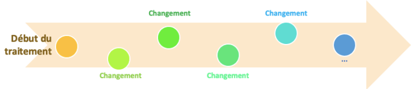 | How do you identify the need to change treatment for a patient? | - Communication: explanations provided, understanding of changes (literacy)<br>- Shared decision-making: treatment chosen, overall vision, short- and long-term treatment management, life plan<br>- Various healthcare professionals consulted: their expectations, positioning, change of professional |
| **<u>Topic n°4 :</u>** **The patiente-environnement relationship** | wooclap | *In your opinion, what socio-environmental factors have a significant influence on the patient and treatment choices?* | - Communication<br>- Sources of information requested<br>- Social environment: relationship with partner, family life, caregiver/support person?<br>- Opinion of family and friends regarding treatment: importance given?<br>- Professional environment?<br>- Role of peers |
